# Knowledge, Skills, and Triage Practices in Emergency Nurses in Mafraq

**DOI:** 10.64898/2026.02.17.26346462

**Authors:** Malek Alrfooh, Iman Eladjaoui

**Affiliations:** Department of Public Health, Ministry of Health, Amman 11118 Jordan.; Department of family medicine (specialist), Ministry of Health, Amman 11118 Jordan.

## Abstract

Emergency nursing is essential to healthcare systems worldwide. Triage plays a pivotal role in emergency nursing, prioritizing patients based on the urgency of their medical condition and focusing on rapid assessment and prioritization of patient care according to their condition and its severity. In the emergency department, the triage nurse assesses vital signs and gathers information from the patient to determine the severity of their condition. This aims to provide appropriate medical intervention quickly for life-threatening cases and minimize waiting times for less critical cases, thus contributing to the efficient allocation of scarce resources. Our study aimed to evaluate the triage knowledge, skills, and practices of emergency nurses in Mafraq, Jordan.

**Methods:** A cross-sectional study used a previously validated questionnaire. Fifty emergency nurses from two public and one private hospital in Mafraq participated. We collected data through an online survey then analyzed in SPSS.

**Results:** 92% of nurses had sufficient triage knowledge (≥60%), while 14% exhibited deficient triage skills (<60%) and 86% had moderate skills (60-80%). Regarding practices, 32% rated as “poor” (<60%) and 68% as “adequate” (>60%). Length working in emergency, hospital type significantly related to nurses’ triage knowledge, skills, and practices.

**Conclusion:** The study underscores continual training’s, simulation programs’ and mentorship’s importance for enhancing emergency nurses’ triage knowledge, skills, especially in rural settings. Implementing clear triage protocols, continuous support and integrating triage competencies into curricula are recommended to improve overall triage competency

## Introduction

Emergency nursing is integral to healthcare systems worldwide. Nurses are responsible for initial patient assessment and care in emergency departments. Triage, prioritizing patients according to medical urgency, forms the backbone of emergency nursing. Through triage, providers efficiently allocate scarce resources and proportion treatment urgency. When facing critical illness, emergency nurses must expertly triage patients to receive life-saving care without delay. However, correctly identifying less dire presentations proves equally important to relieving suffering and ensuring proper subsequent care (Aloyce et al., 2014).

Effective triage practices are essential for ensuring that patients receive timely and appropriate care, particularly in resource-constrained settings (Phukubye et al., 2021).

In Jordan, emergency nurses exhibited a moderate grasp of triage-related knowledge and competencies in Amman hospitals. Their learning encompassed topics central to prioritizing patients according to acuity, though several influences imparted varying command. Level of education, longevity on the job, and availability of specialized preparation all affected nurses knowledge (Malak et al., 2022). Comparatively, a recent South African work by Phukubye et al (2021) revealed that emergency nurses in rural hospitals had limited knowledge and practical skills related to triage. Duko et al. (2019) also reported that emergency nurses had suboptimal triage knowledge and skills, with significant gaps in their understanding of triage principles and their ability to accurately prioritize patients.

Several studies have explored the factors that influence the triage practices of emergency nurses. In a study conducted in Jordan, AlShatarat et al. (2022) found that the nurses’ level of education, years of experience, and access to triage training were positively associated with their triage knowledge and practice.

The researchers also identified organizational factors, such as staffing levels and workload, as important determinants of triage performance.

Similarly, a study in Nigeria by Nneoma et al. (2022) reported that emergency nurses’ perceived skills in triage were influenced by their level of education, years of experience, and access to triage-specific training, emphasizing the need for targeted interventions to enhance the nurses’ triage knowledge and skills. In Iran, Reisi et al. (2018) also found that the level of awareness of triage principles among emergency department nurses was generally low, with significant variations across different hospitals. Highlighting the importance of standardized triage protocols and ongoing training to ensure consistent and effective triage practices. Similarly, Nneoma et al. (2022) emphasized the need for regular triage-specific training, the establishment of triage committees, and the implementation of quality assurance measures to ensure the consistent and effective application of triage practices. Bahlibi et al., (2022) also stated that training significantly improved the nurses’ knowledge application and triage practices.

Differences in results among different demographic variables raises the need for more research in this area. The previous studies also highlight the importance of the issue and its probable effects on patient outcomes specially in critical cases. It is obvious too that there is scarcity of research in Jordan regarding this issue in rural areas.

The objective of this work was to study the levels and associated factors of knowledge, skills, and practices of triage among emergency nurses in Mafraq, an eastern Jordanian city, and extend conclusions on the levels of knowledge among this population.

## Methods

The study utilized a cross sectional design utilizing a previously validated questionnaire mentioned below.

In the present study, the sample was made up of all emergency nurses who were employed under the triage category in the Emergency Department across two governmental hospitals and one private hospital in Mafraq. The Initial sample size was calculated using alpha value of 0.50, power = 0.80, moderate effect size of 0.30, and 2 of freedom on chi square test. The initial required sample size was 45, though it was increased to 50 in consideration of missing data. The study included registered nurses having not less than 6 months of exposure in the emergency department and were willing to take part in the study. Nurses working in administrative positions were excluded.

Ethical approval was gained from the Ministry of health. Data was collected through a cross-sectional online Google form survey, utilizing a self-administered questionnaire. The researcher met the nurses in emergency units and invited them to participate in the study. The researcher offered guidance on how to approach the participants via sharing the link with the head nurses on whatsapp and answering the research.

Participants were assured of anonymity and that their identifying information were not collected. All data collected was treated as confidential and used solely for research purposes. The sampling process was conducted online, with the questionnaire distributed via a Google form to all associated nurses.

The questionnaire included a part for collecting demographic data, including age, gender, level of education, length of employment in the emergency department, length of employment in triage, completion of triage training, and other training courses.

The Triage Knowledge Questionnaire (TKQ), created by Fathoni et al (2013) was used for assessment. It contains 13 questions created to measure one’s understanding of triage. Each question earned one point for a right answer and zero is given for a wrong one. The total score of the questionnaire is between 0 and 13. Total score of less than 60% indicates a lack of proper knowledge of triage. A score of 60% or more shows adequate triage knowledge. TKQ had high validity and reliability, which can be realized in a test-retest correlation of 0.99 (Fathoni et al., 2013), (Malak et al., 2022)

Triage Skills Questionnaire developed by Fathoni et al (2013) was used for assessment purposes. It comprises 37 statement items used for evaluation of technologies in triage. The tool includes three aspects: quick assessment comprising 27 items, determination of degree of illness or patient classification (4 items), and patient allocation (6 items). The responses were rated on 5-scale Likert scale: 1 means much room for improvement, 2 is bad, 3 is fair, 4 is good, and 5 is very good. The questionnaire had a total score ranging from 37 to 185 and the score was converted into percentage and categorized as follows: conducting poorer triage if the score was below 60%, fair if the score ranged between 60% to 80%, and good if the score exceeded 80%.

Evaluating methods of prioritizing and categorizing patients based on their severity of illness. Was done using Triage Skill Observational Checklist by Aloyce et al (2014). Ten items were used to validate categorization in Emergency Department operational practices, and the tools were used in the form of direct observation of triage methods used by nurses in emergency department. Each aspect had yes and no response with a total score below 13 being considered poor practices, while 13 above was good practices. The item validity and reliability of the item were satisfactory.

The internal reliability of this questionnaire in English language was assessed by Malak et al (2022) (Cronbach’s alpha: 0.85, and 0.95, respectively).

Statistical analysis was performed using SPSS version 23. Descriptive statistics were used to describe the sociodemographic characters. Chi-square test of significance test was utilized for measuring the association between the independent variables (gender, educational level, training experience, training courses in triage, duration of training triage course, and working experience) and dependent variables (triage knowledge, skill, and practices).

Informed consent was obtained from all participants prior to their inclusion in the study. Participation was voluntary, and all participants were informed about the study objectives, procedures, potential risks, and their right to withdraw at any time without consequences. The study was conducted in accordance with the ethical principles of the Declaration of Helsinki for research involving human participants. Ethical approval for this study was obtained from the Research Ethics Committee of the Jordanian Ministry of Health (MOH/REC/2024/2)

## Results

Table 1 reveals demographic and professional information about a group of emergency nurses. 60% of nurses are males, while 40% are females. 50% of nurses are 30-33 years old, only 16% were older than 33, and 14% between 22-25, 68% of nurses work in governmental hospital, and 32% work in private hospital. 86% of the nurses have a postgraduate diploma, while 14% have higher education.

**Table 1:** Demographic characteristics of the participants (N = 50).

|  | Frequency | Percent% |
| --- | --- | --- |
| **Gender:** |  |  |
| Male | 30 | 60 |
| Female | 20 | 40 |
| **Where are you currently working/ hospital?** |  |  |
| 22-25 | 7 | 14 |
| 26-29 | 10 | 20 |
| 30-33 | 25 | 50 |
| >33 | 8 | 16 |
| **Where are you currently working/ hospital?** |  |  |
| Governmental Hospital | 34 | 68 |
| Private Hospital | 16 | 32 |
| **What is your level of nursing education?** |  |  |
| Post graduate diploma | 43 | 86 |
| Higher Education: | 7 | 14 |
| **How long have you been working in the emergency department?** |  |  |
| Below 1 year | 5 | 10 |
| Between 1 to 3 years | 14 | 28 |
| Between 3 to 5 years | 13 | 26 |
| Between 5 to 10 years | 10 | 20 |
| Others | 8 | 16 |
| **Have you ever been trained on emergency nursing care?** |  |  |
| yes | 50 | 100 |
| **If YES on question 6, did the training include how to triage patients?** |  |  |
| yes | 50 | 100 |
| **Have you been working in the triage unit?** |  |  |
| yes | 50 | 100 |

28% of nurses have 1-3 years of experience, 26% have 3-5, only 10% have less than 1 year of experience. All nurses received training for emergency nursing care, and all of those have also been trained on the job for triage of patients.

Table 2 shows the results of the chi test of significance. With a value of 19.251 with a p value of implies that there is a significant relationship regarding the gender. This suggests a difference in nursing practice, and skills between the males and their female counterparts.

**Table 2:** Results of Chi-square test and significance of demographic variables with triage knowledge, skills, and practices.

|  | knowledge |  | practice |  | skill |  |
| --- | --- | --- | --- | --- | --- | --- |
|  | chi | p-value | chi | p-value | chi | p-value |
| Gender | .181 | 0.67 | 19.251 | 0 | 12.209 | 0 |
| Where are you currently working/ hospital? | 26.708 | 0.00 | 16.001 | 0.001 | 15.828 | 0.001 |
| Where are you currently working/ hospital? | 0.647 | 0.42 | 8.517 | 0.004 | 0.441 | 0.507 |
| What is your level of nursing education? | 0.708 | 0.40 | 0.107 | 0.744 | 1.325 | 0.25 |
| How long have you been working in the emergency department? | 21.079 | 0.00 | 18.662 | 0.001 | 16.303 | 0.003 |

There was also a significant relationship at p value of 0.0001 of current work place and nursing knowledge, practice and skills. However, regarding nursing education, no significant difference was found.

Regarding working in the emergency department, a significant relationship is clear with a p value of 0.003 in each as chi value of 21.079, 18.662a and 16.303 in nursing knowledge, practice, and skills respectively suggest very strong relationship with emergency depart.

The results revealed that the vast majority of nurses (92%) had sufficient triage knowledge (≥60%).

However, 14% exhibited poor triage skills (<60%), while the remaining 86% had moderate skills (60-80%). When it came to triage practices, 32% were rated as “bad” (<60%), while 68% were considered “good” (>60%) (Table 3).

**Table 3:** Levels of triage knowledge, skills, and practices among emergency nurses.

| variable | N(%) |
| --- | --- |
| Triage knowledge |  |
| Insufficient (<60 %) | 4(8%) |
| Sufficient (≥60 %) | 46(92%) |
| Triage skills |  |
| Poor (<60 %) | 7(14) |
| Moderate (60–80 %) | 43(86%) |
| Triage Practices |  |
| Bad (<60 %) | 16(32%) |
| Good (>60 %) | 34(68%) |

## Discussion

Educational interventions have been shown to significantly improve nurses’ knowledge, attitudes, and practices related to triage in emergency departments (Atigo & Yousif, 2021). Our results showed a significant relationship between the length of time working in the emergency department and the nurses’ triage knowledge, skills, and practices. This suggests that hands-on experience in the emergency setting is an important factor in developing and maintaining triage expertise. The search results indicate that all the nurses had received training in emergency nursing care, including specific training on triage. This emphasis on triage training likely contributed to the high percentage (92%) of nurses demonstrating sufficient triage knowledge. However, the results also show that only 86% had moderate triage skills and 68% had good triage practices, suggesting that knowledge alone is not enough, and practical skills training is crucial.

This study found a significant relationship between the type of hospital (governmental vs. private) and the nurses’ triage knowledge, skills, and practices. This suggests that the work environment, resources, and organizational support available in different healthcare settings can impact the development and application of triage competencies.

While gender did not show a significant difference in triage knowledge, the results indicate a significant difference in triage practices and skills between male and female nurses. This could be due to factors such as differences in confidence, decision-making styles, or opportunities for hands-on triage experience.Comparing these findings to previous literature, the study aligns with the importance of experience in the emergency department for triage competency, as highlighted by Malak et al. (2022) and AlShatarat et al. (2022). The emphasis on training for emergency nursing care and triage is consistent with the literature, emphasizing the need for ongoing education and skill development in this critical area of nursing practice. Our results showed higher level of knowledge compared to (Twagirayezu et al., 2021) and (Duko et al., 2019) who reported low level of knowledge among nurses in Rawanda and Ethiopia respectively. Differences in Results among different countries can be attributed to different methods used, different settings and availability of resources.

When compared to the results of other studies such as Phukubye et al. (2021), Bahlibi et al. (2022), and Duko et al. (2019), the findings of this study are consistent with the emphasis on training and experience in the emergency department as key factors influencing triage knowledge, skills, and practices among nurses. The high percentage of nurses with sufficient triage knowledge aligns with the positive impact of training programs highlighted in previous research. The significant relationship between workplace and triage knowledge, practice, and skills emphasizes the importance of the work environment in shaping nurses’ competencies in triage. It is recommended to implement structured simulation based training programs, develop clear triage protocols, provision of ongoing support and supervision for emergency nurses, and integration of triage competencies into nursing curricula to improve the knowledge and practices of emergency nurses in rural settings (Phukubye et al., 2021).

Bruce and his colleagues (2018) explored the utility of simulation as a means to boost triage expertise for emergency room nurses. Through simulation, nurses were offered chances to rehearse and refine their abilities. According to their findings, simulation proved good at enhancing nurses’ clinical decision-making and self-assurance when facing triage tasks. Rankin et al (2013), led a study examining the potency of web-based triage learning. Most participants at 74% noted simulations improved their triage proficiency. The researchers emphasized simulations’ knack for cultivating judgment and composure in nurses grappling with triage determinations. Overall, their work highlighted simulation’s power as an instrument for optimizing triage acumen in nursing staff.

## Recommendations

1. Ongoing continuous programs should implement focusing to enhance triage especially for exhibited poor skills. These programs aim to improve decision-making and patient prioritization during include hands-on practice sessions and simulations.
2. Conduct workshops to the practices of rated as “bad.” These emphasize efficient patient flow management, accurate assessment, and effective in high-pressure situations.
3. Develop specialized training modules for with less than 1 year of experience to ensure a solid foundation in practices. These protocols, critical thinking skills, and rapid assessment techniques.
4. Establish mentorship programs where experienced can guide and support those with limited experience in the emergency department. This will facilitate knowledge transfer and skill development, enhancing overall triage competencies.
5. Implement regular skill assessments and performance evaluations to monitor the progress of nurses in triage knowledge, skills, and practices. Feedback from assessments can help tailor training programs to address specific areas of improvement.

## Conclusion

The high percentage of nurses with sufficient triage knowledge in Mafraq is promising, but there is a need to focus on improving triage skills and practices, especially among those with limited experience and lower skill levels. Policy makers should implement targeted training programs, skill development workshops, mentorship initiatives, and regular assessments to enhance the overall quality of triage care provided by emergency nurses.

## Data Availability

All relevant data are within the manuscript and its Supporting Information files.

